# Status of Compassionate Health Care Practice and Associated Factors in Public Hospitals of Addis Ababa from patient’s perspective: cross sectional study

**DOI:** 10.1101/2023.06.06.23291023

**Authors:** Elias Teshome Tadesse, Dawit Regasa Soboka, Gurmesa Tura Debelew

## Abstract

**Background:** Compassionate health care is lauded as a cornerstone of quality healthcare by patients, families, clinicians, and policy makers. Compassionate Care’ within the healthcare setting has received much attention globally, following concerns that healthcare often fails at a fundamental level.

**Objective:** To assess Status of compassionate health care practice and associated factors in public hospitals of Addis Ababa.

**Methods:** Institution based cross-sectional study design was conducted. A structured and prtested interviewer administered questionnaire was used to collect the data from 605 study participants. Study participant was selected using a systematic sampling technique by allocating a proportion sample to each hospitals. The data was entered with Epi data version 3.1 and exported to Statistical Package for Social Sciences version 25.0 for further analysis. Both bivariate and multivariate logistic regression analysis were performed to identify associated factors. P values < 0.05 with 95% confidence level were used to declare statistical significance.

**Result:** The status of compassioante health care practice was found to be 42.6% [(95% CI: 39.0%-47.0%)]. Covariates such as: relatively higher education [AOR = 0.534 (95% CI 0.305-0.934)], Not married [AOR=0.561 (95% CI 0.330-0.955)], government employee [AOR=2.178 (95% CI 1.071-4.430)], Room accommodation comfortability [AOR=0.309 (95% CI 0.178-0.535)], and Cleanness of toilet [AOR=2.158 (95% CI 1.326-3.513)] had statistically significant association with compassionate health care practice.

**Conclusion:** The status of compassionate health care practice was found to be low compared to similar study conducted with in and out of Ethiopia. Compassionate health care practice was significantly associated with educational s, occupation, marital status, cleanness of toilet, and room accommodation comfortability. Therefore, health workers, Hospitals and Addis Ababa health bureau should understand these weak areas and plan for a better service delivery to compassionate care.

## Background

Compassion is a feeling of deep sympathy and sorrow for the suffering of others accompanied by a strong desire to alleviate the suffering (1). Compassionate Care’ inside the healthcare setting has gotten much attention globally, taking after concerns that healthcare often falls flat at a fundamental level (2). In spite of the expanding scope and modernity of healthcare, the tremendous assets given to it and the center on advancement; it is still falling flat at a crucial level. Caring and compassion, the essentials of care delivery, and the human aspects that characterize it appear to be beneath strain. The parts of caring, comfort and compassion have been supplanted with a basic center on pathways, tasks and documentation though it is paramount important and indispensable (3).

A number of patients encounter de-humanizing and impersonal treatment. This crisis within the health system gets to be damaging not as it were for patients but too for their families, health professionals and the health system itself (4). The current depersonalize and dehumanization of healthcare regularly take off patients feeling like ‘the kidney in Room 5,’ or ‘the liver in Room 10’, instead of persons, unique individuals with a special story of sickness (5).

A study done in South Africa concluded that patient fulfillment may be a essential pointer of impartial quality of care. In reality, satisfied patients are likely to display favorable behavioral intentions, which are advantageous to the healthcare provider’s long-term victory. Be that as it may, one of the major obstructions to way better health care for much of the population in developing countries, is lack of access to even fundamental health service (6).

According to numerous studies the recognized barriers to execute and bolster compassionate care are time constraints, work load and staffing levels, Resistance to change, Lack of organizational support, Lack of inclusion of front-line staff into care planning and Lack of resources (3, 7, 8).

Aiding health professionals’ to become compassionate and respectful professionals remains a major challenge for the health care. However the modern national Health Sector Transformation Plan (HSTP) created in 2015 has underlined the significance of making compassionate health workforce as a major pillar to improve the quality of health care services. This has driven to a reestablished focus on how to improve the health workforce execution including responsiveness, timeliness and persistent centeredness of healthcare services (1, 3). Despite the problem, little research has been conducted in Ethiopia with regard to the status of compassionate care. Therefore this study aimed at assessing the status of compassionate health care practice and associated factors from patients perspective in public Hospitals of Addis Ababa.

## Methods

### Study area

The study was conducted in Addis Ababa, the capital of Ethiopia. The city covers 527 square kilometers and with estimated population of 7,823,600 in 2019 (9). In Addis Ababa there are about 38 hospitals comprising about 12 governmental and 26 non-governmental. The study was conducted in six public hospitals.

### Study design and period

Institutional based cross-sectional study was conducted from July 27-Sep 09, 2020.

### Population

Patients attending in public hospitals of Addis Ababa

### Sample size determination

The sample size was determined using single population formula. The sample size calculation assumed the proportion (P) estimated level of compassionate care 45.7% study done in Addis Ababa at Tikure Anbessa Specialized Hospital (10). Adding non response rate 10%, margin of error 5%, 1.5 design effect and considering 95% confidence interval the final sample size was 629.

### Sampling procedure

Out of 12 public hospitals in Addis Ababa six hospitals were selected by using computer generated simple random sampling. Each selected hospital was stratified into outpatient and inpatient strata. The allocation of the sample to hospitals was made proportionally based on the average number of patients who received services at each hospital in the month preceding the data collection period. Yekatit 12 Medical College General Hospital 165 outpatient and 9 inpatients, Ras Desta Damtew Memorial General Hospital 55 outpatients and 7 inpatients, Zewuditu Memorial General Hospital 123 outpatients and 5 inpatients, Kidus Paulos Specialized Referral Hospital 123 outpatients and 19 inpatients, Gandhi Memorial General Hospital 73 outpatients and 4 inpatients and Dagmawi Minilik Referral Hospital 44 outpatients and 14 inpatients. Individual participants in each of the hospitals was selected by systematic random sampling during the data collection period until the required sample size at each hospital obtained.

### Operational definitions

Compassionate care practice: to say that health care provider has demonstrated compassionate care study participants has to rate 9 or 10 to the elements of compassionate care. Since the validation analysis have shown that the Schwartz Center Compassionate Care Scale (SCCCS) has 10 items with a two factor structure the minimum cut point to say that compassionate care is demonstrated by the health care provider is 90 point (10).

### Data collection tool

The questionnaires for data collection were initially prepared in English, and translated into the local language (Amharic) and back into English to check for consistency with language experts. Participants were interviewed during discharge for hospitalized patients and at exit for OPD patients. By using a pre-tested structured Amharic version questionnaire. The tool was a validated tool for assessing compassionate care which was the SCCCS. The other included questions in the questionnaire were prepared by reviewing different other related works of literature and variables identified to be measured.

### Data collection procedure

A total of 2 data collectors and 1 supervisor were recruited for data collection procedure. The data collectors and supervisor were non-staff members. The questionnaires were filled by direct face-to-face interview. Clients were interviewed during discharge for hospitalized patients and at exit for OPD patients. The clients were interviewed outside the service room far away from employees. The data collectors and supervisor were trained a week ahead of the actual data collection period on data collection process to standardize interviews and reduce interviewer biases.

### Data analysis

First the collected data was checked manually for completeness and any incomplete or misfiled questions. The data was cleaned and stored for consistency, entered into Epi Data version 3.1 software and the accuracy of the data entry was checked by double data entry. Any errors identified during data entry was corrected by reviewing the original completed questionnaire. Then exported to statistical package for social sciences (SPSS) version 25.0 software for analysis. Univariate analysis like simple frequencies tables, percentages, mean, standard deviation, bar chart, and pie chart was used. Initially, bivariate logistic regression was carried out to see the association of each of the independent variables with the outcome variable. Thereafter, the multivariable logistic regression method was used. The variables that were not significant in the bivariate logistic regression were not considered in the multiple regression analysis. P-Value of < 0.05 and 95% confidence level was used as a difference of statistical significance. Finally, results were compiled and presented using tables, graphs, and text.

## Result

### Socio-demographic and socio-economic status of the participants

In this study, 605 patients have participated, which yield 96.2% of response rate. The mean age of the participants was 33.62 ± 11.66 SD, with a range of 18-101 years. Among them, 406 (67.1%) were female, 501 (82.8%) came from Addis Ababa, 577 (95.4%) urban resident, 410 (67.8%) married, 227 (37.5%) were grade 9-12, 128 (21.2%) housewife. Wealth index analysis shows, 164 (27.2%) of the participants were found in poorest wealth quintile (below 20^th^ quintile) and 120 (19.9%) in the richest wealth quintile (above 80^th^ quintile). See table 1

**Table 1.**
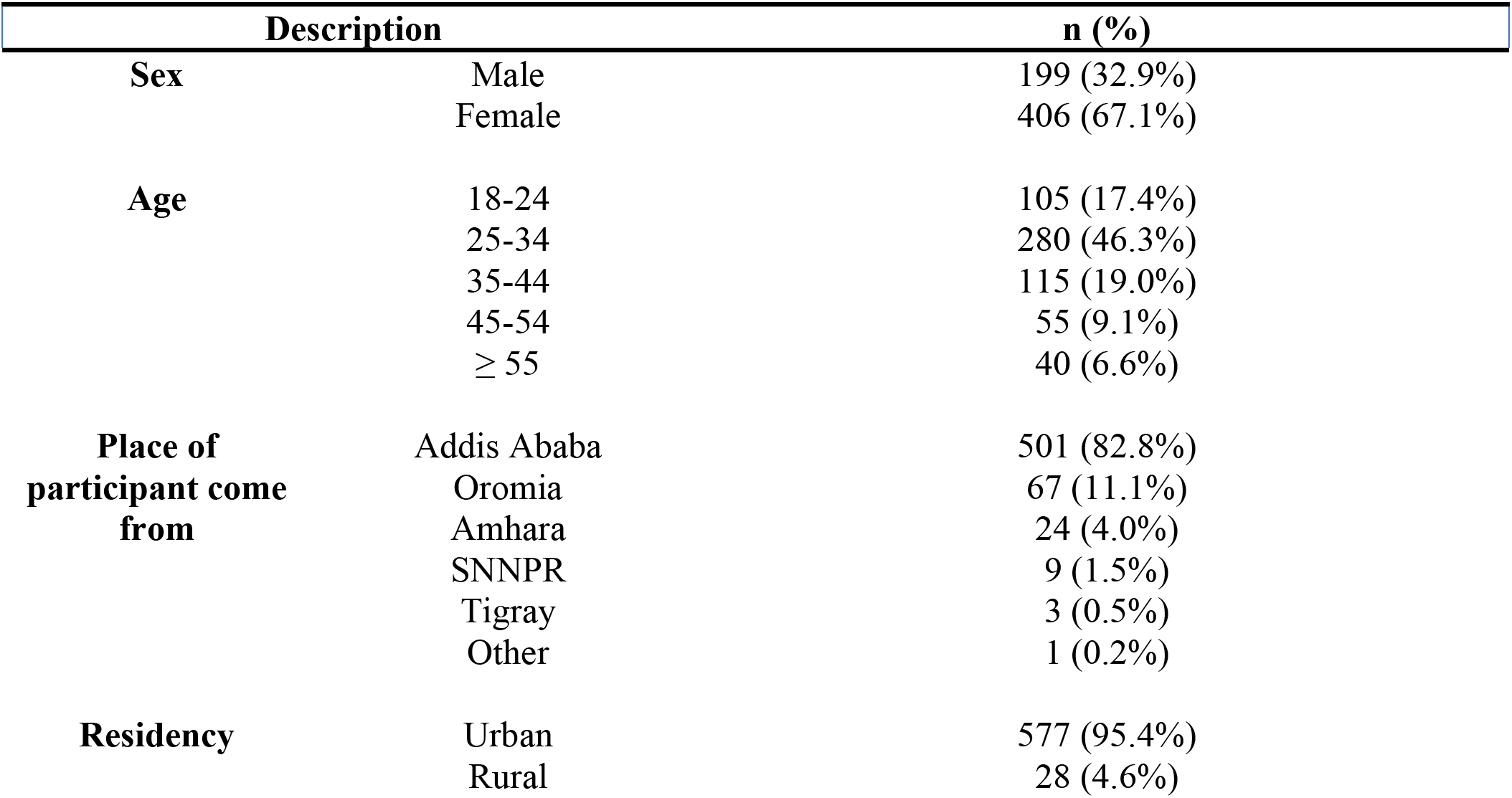
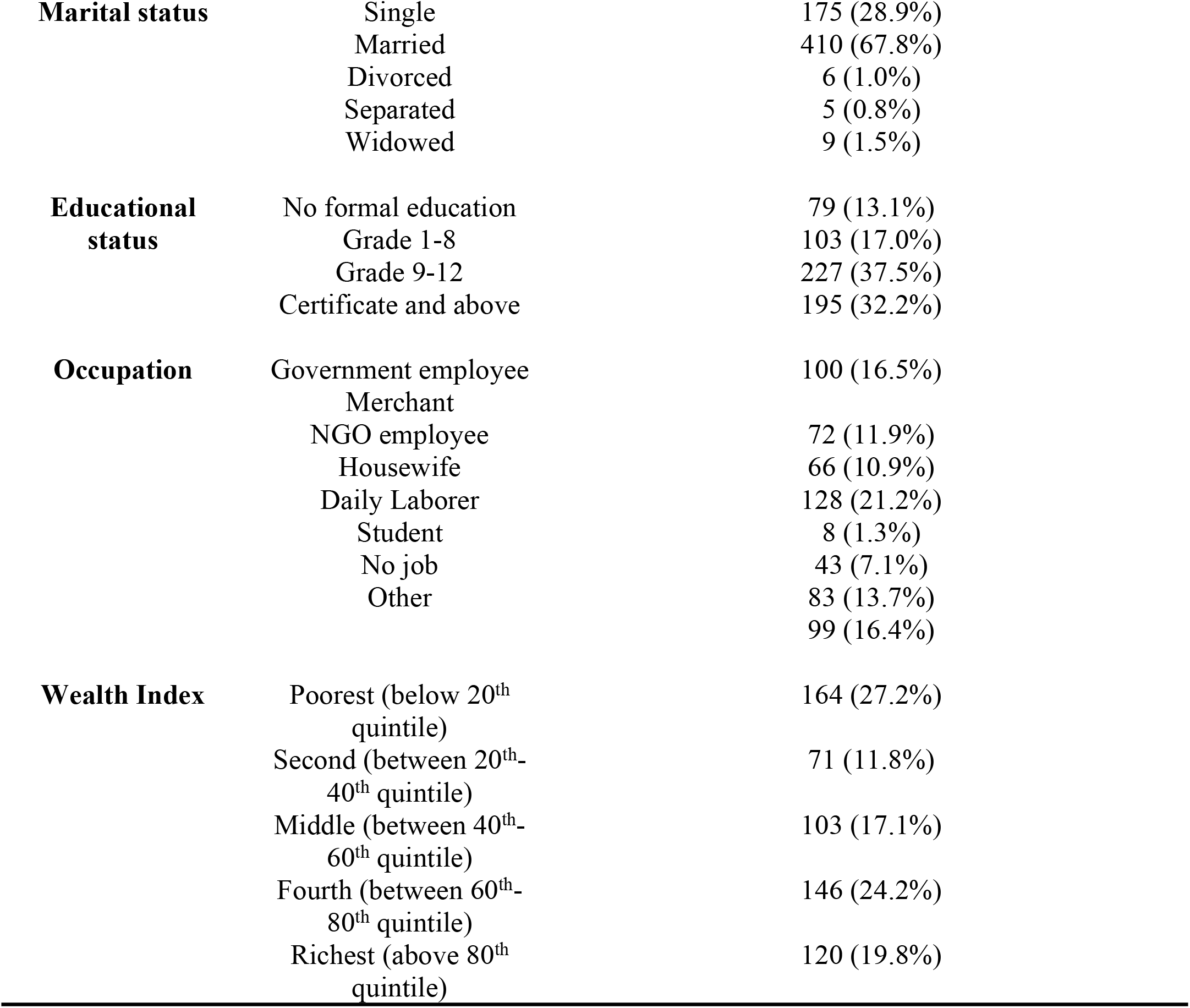
Socio-demographic and socio-economic status of the participants on compassionate health care practice and associated factors, 2020. (n=605)

### Participants medical history

The study participants were included from OPD and inpatient department. Among this majority 544 (89.92%) of participants were from outpatient department. The mean number of visits for outpatient was 11.44 ± 20.86 SD with the range of first time to ten plus years. The mean day of hospitalization was 11.20 ± 15.00 SD with the range of 1-80 days. The mean number of hospitalizations was 6.49 ± 17.60 SD with the range of one to more than ten times.

### Health care environment

Majority 551 (91.1%) of participants thinks the ward or outpatient department was clean, 430 (71.1%) of participants thinks the accommodation was comfortable, 520 (86.0%) of participants do not thinks the bed or exam was clean. In addition to this 277 (45.8%) of participants thinks the toilet was not clean.

### Compassionate health care practice

Compassionate health care practice was measured from patient perspective using SCCCS, by rating a brand score of 1-10. Less than half 258 (42.6%) with (95% CI: 39.0%-47%) participants reported compassionate care was demonstrated. From compassionate care components “Treat you as a person, not just a disease” gets the highest percent (77.4%). Whereas “Communicate test results in a timely and sensitive manner” gets the least percentage (35.4%). See table 2

**Table 2.**
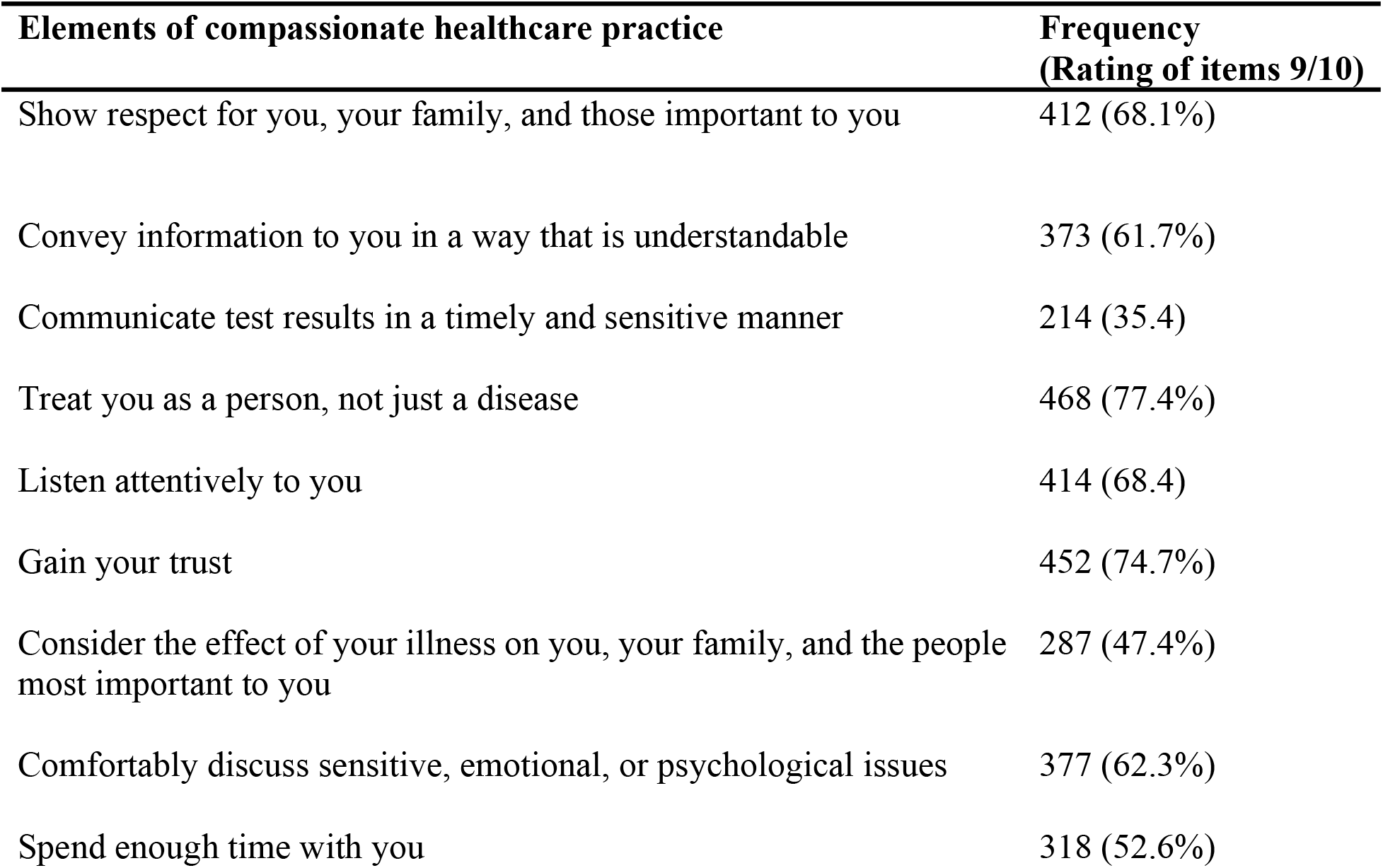
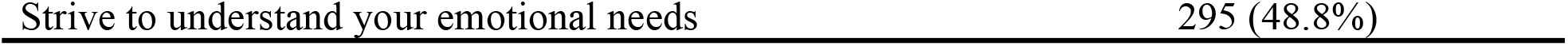
Participants rating items for compassionate health care practice as health care providers in public hospitals of Addis Ababa, Ethiopia, 2020, demonstrate it. (n=605).

The overall compassionate health care practice in Public Hospitals of Addis Ababa was found to be 258 (42.6%) with (95% CI: 39.0%-47%). See figure 1

**Figure 1.**
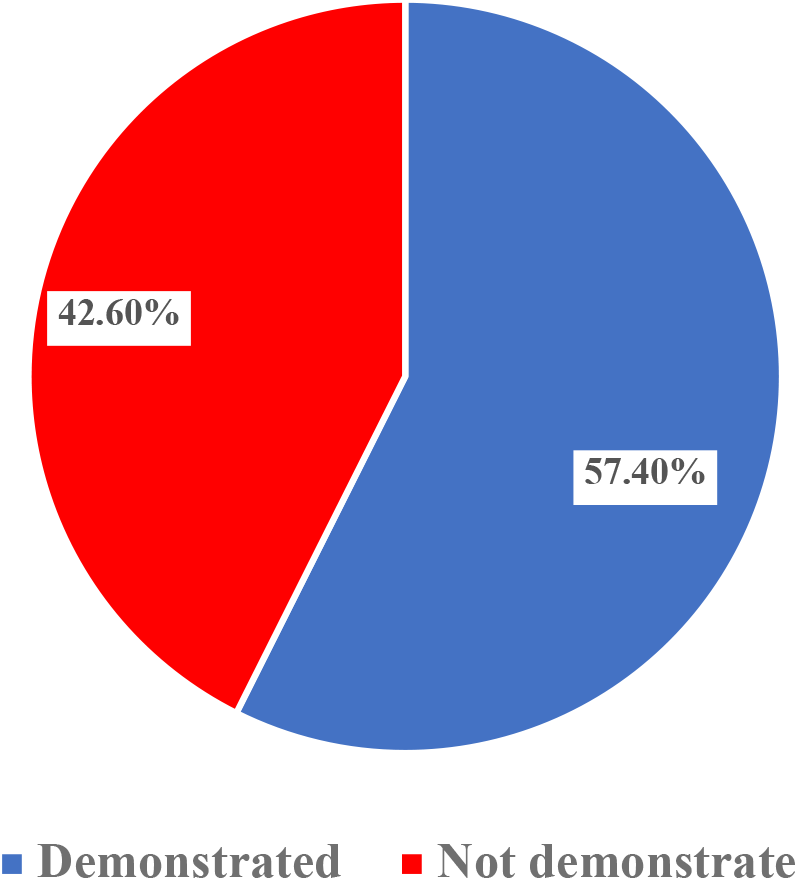
Status of compassionate health care practice from patient perspective in public hospitals of Addis Ababa, Ethiopia, 2020. (n=605)

### Facctors associated with compassionated health care practice

In multivariable logistic regression analysis: marital status, educational status, occupation, room accommodation comfortability, and toilet cleanness were significantly associated with reporting compassionate health care practice. Not married participants were about 44% less likely to report compassionate health care practice than their married counterparts [AOR=0.561 (95% CI 0.330-0.955)]. Those who had certificate and above (relatively higher education) were about 47% less likely to report compassionate health care practice compared to grade 9-12 (relatively lower education) [AOR = 0.534 (95% CI 0.305-0.934)]. Government employee were more than 2.1 times to report compassionate health care practice as compared to housewife [AOR=2.178 (95% CI (1.071-4.430)]. Room accommodation comfortable ability was significantly associated with reporting compassionate health care practice. Participants were 69% less likely to report compassionate health care practice when the room accommodation was not comfortable [AOR=0.309 (95% CI 0.178-0.535)]. Cleanness of toilet facility had positive association with reporting compassionate health care practice. Participants who thinks toilet room was clean and didn’t see the toilet facility were more than 2 and 4 fold to report compassionate health care practice [AOR=2.158 (95% CI 1.326-3.513)] and [AOR=4.269 (95% CI 2.458-7.414)] respectively. See table 3.

**Table 3.**
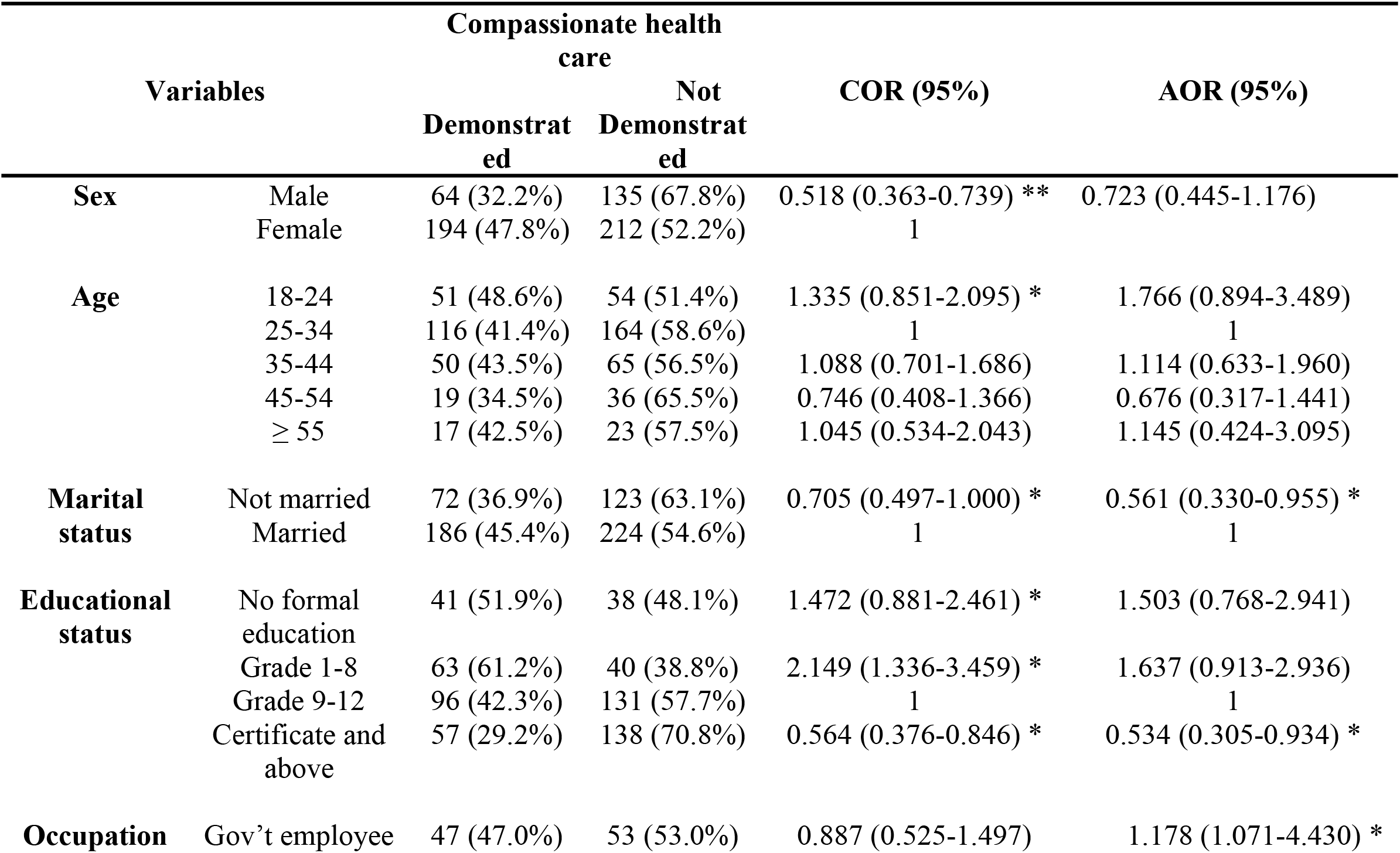
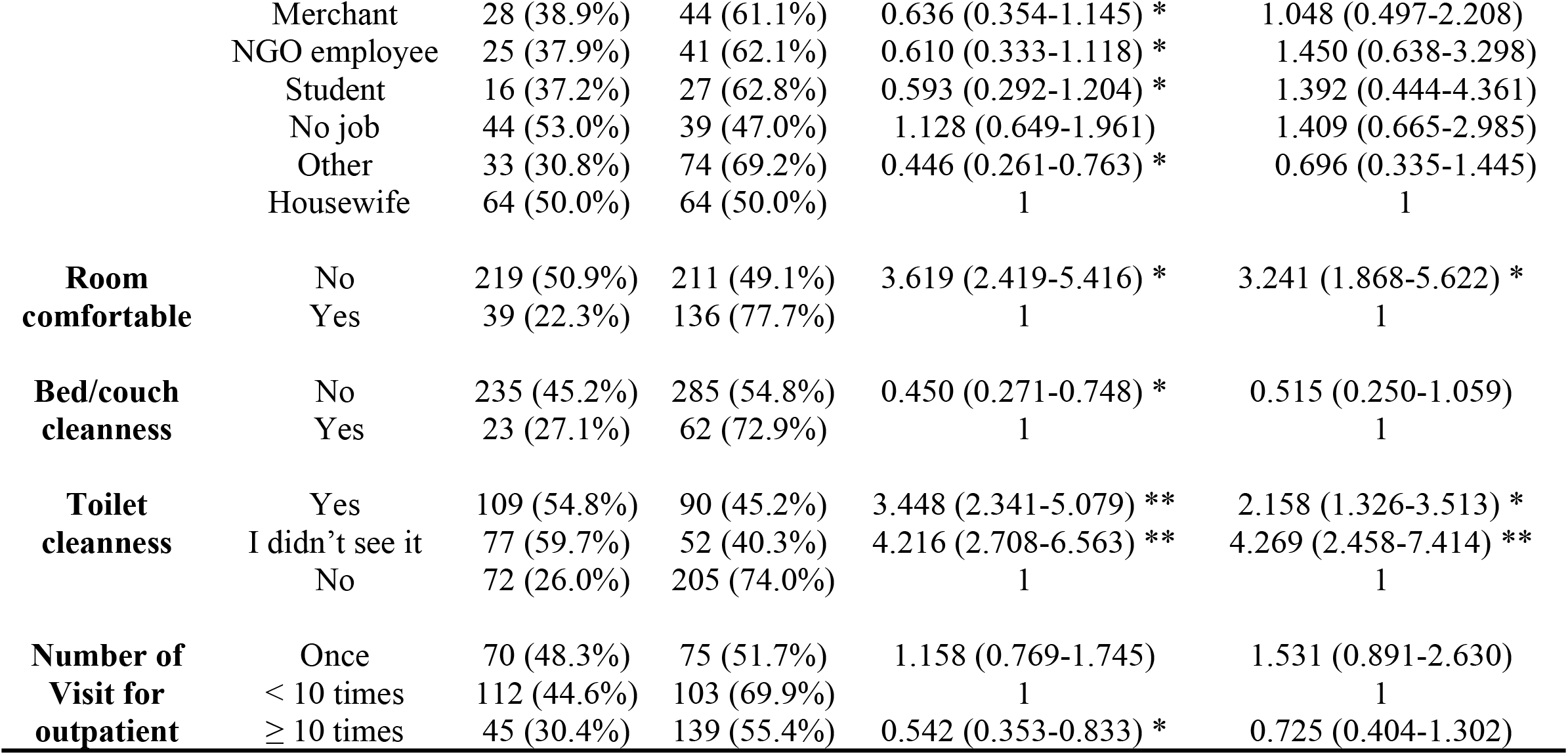
Factors associated with compassionate health care practice in public hospital of Addis Ababa, multivariable logistic regression analysis, 2020. (n=605)

## Discussion

The overall compassionate health care practice in Public Hospitals of Addis Ababa was found to be 42.6% [(95% CI: 39.0%-47.0%)]. From compassionate care components “Treat you as a person, not just a disease” gets the highest percent (77.4%). Whereas “Communicate test results in a timely and sensitive manner” gets the least percentage (35.4%). This lower than a study done in USA 53% (11). This difference could be due to the emphasis given to compassionate health care practice is much lower than a country like USA. due to the emphasis given to compassionate health care practice is much lower than a country like USA. Moreover distinction in health care providers awareness level on compassionate health care might play a huge part.

A study was conducted in Saudi Arabia to determine the level of awareness of patients’ rights among hospitalized patients. According to this study 75.4% patients believed that they receive compassionate and respectful care (12). Which is better than the findings of the current study (42.6%). The reason for the difference may be attributed to the type of tool used and the approach of data collection.

A study was done in Tigray region to asess the status of compassionate and resprectful care from patient prespective. According to this study 55% patients had good experince (3). Again this is also better than current study (42.6%). This variation might be due to difference in sample size, tool and study area.

Current study is lower than a study done at Tikur Anbesa Specilized Hospital on oncology patients which (45.7%) even though both used the same tool (10). This difference might be variation in sample size and current study included 6 different public hospitals instade of only one.

In multivariabl logistic regression analysis, educational status shows negative association with reporting compassionate health care practice. Those who had certificate and above (relatively higher education) were about 47% less likely to report compassionate health care practice compared to grade 9-12 (relatively lower education). This is supported by study conducted in USA, respondents with a college degree or higher were less likely to report positive patient exprince than those with less education (13). Also inpatient study in Ghana indicated patients with high formal education were less satisfied than their counterparts (14). Smilarly, the study conduceted at Debra Marko (15), indicated that those who were unable to read and write were about 2 times more likely satisfied than those who were literate.

This may be the result of patients with higher education being able to get to information about the obligations of the health care providers. On the off chance that these obligations are not carried out well, they may not be satisfied.

Occupation status of participants had a positive association with reporting compassionate health care practice. Government employee were more than 2.1 times to report compassionate health care practice as compared to housewife counterpart. This contradict from study conducted in Felegehiwot hospital; showed employees were 58% less likely to satisfy than non employees (16). This might be due to government employee knows the bureaucracy in government institutions and know what kind of service they might get compared to their counterparts.

Marital status had negative association with reporting compassionate health care practice. Not married participants were about 44% less likely to report compassionate health care practice than their married counterparts. This finding was consistent with a study done in Northwest Ethiopia (16) and Dessie Referral Hospital (17).

The room accommodation comfortablity was significantly associated with reporting compassionate health care practice. Participants were 69% less likely to report compassionate health care practice when the room accommodation was not comfortable. The reason for this might be, when the room accomedation is comfortable patient’s or client’s will feel more secure to talk freely and take physical examination.

Cleanness of toilet facility had positive association with reporting compassionate health care practice. Participants who thinks toilet room was clean and didn’t see the toilet facility were more than 2 and 4 fold to report compassionate health care practice respectively. This was line with study conducted in Yekatit 12 Medical College (18) and Wolita-Sodo University Teaching Hospital (19). This might be attributed to COVID 19, since the start of the pandemic more attention was given for sanitation as a part of one prevention method.

## Conclusion

The overall status of compassionate health care practice from patient perspective was found to be low as compared to different literatures. Compassionate health care practice was significantly associated with educational status, occupation,marital status, cleanness of toilet, and room accommodation comfort ability. Therefore Health care providers ought to create the aptitudes of compassionate care as such of the scientific knowledge of medicine. And Hospitals should make extra effort to make their environment clean and patient centered.

## Data Availability

All relevant data are within the manuscript and its Supporting Information files.

## Abbrevations

AOR: Adjusted odds ratio
CI: Confidence interval
HSTP: Health Sector Transformation Plan
SCCCS: Schwartz Center Compassionate Care Scale

## Acknowledgements

The authors are grateful for the data collectors and study participants.

## Availability of data and materials

The datasets used and/or analyzed during the current study available from the corresponding author on reasonable request.

## Authors’ contributions

ET coined the reaserch idea, ETT, DRS, and GTD designed the method. ETT supervised the data collection. ETT, DRS and GTD analysed the data. ETT wrote the initial draft while DRS, GTD and ETT further enriched the initial draft and discussed the interpretation and the implication of the research output.

## Ethics approval and consent to participate

Ethical clearance was obtained from Institution Review Board (IRB) of Jimma University. Ethical clearance was given to secure permission of access to the hospitals included in the study. The study posed a low or not more than a minimal risk to the study participants. Also, the study did not involve any invasive procedures. Accordingly, after the objective of the study was explained, verbal informed consent was obtained from all participants over the age of 18 years. Moreover, the confidentiality of information was guaranteed by using code numbers rather than personal identifiers and by keeping the data locked.

## Consent for publication

Not applicable.

